# Long-term efficacy and safety of single fecal microbiota transplantation for recurrent active ulcerative colitis

**DOI:** 10.1101/2020.03.16.20022293

**Authors:** Haiming Fang, Lian Fu, Xuejun Li, Jiajia Wang, Kangwei Xiong, Yuan Su, Lijiu Zhang

## Abstract

**Aims:** To assess the long-term safety and efficacy of single fecal microbiota transplantation (FMT) for recurrent ulcerative colitis (UC).

**Methods:** 20 UC patients were randomly divided into single FMT (n=10) and standard of care (SOC) (n=10) group. Patients in FMT group were just treated with single fresh FMT. Patients in SOC group with mild to moderate UC were treated with mesalazine, those with severe UC were given corticosteroids-induced remission, mesalazine maintenance treatment. The primary endpoint was clinical and mucosal remission at week 8. The second endpoint was the maintenance of clinical and mucosal remission, and possible adverse events during the long term follow up (12 to 24 months).

**Results:** 90% (9/10) patients in FMT group and50% (5 /10)in SOC group could achieve primary endpoint at week-8.After 12 months of follow-up, 66.7% (6/9) FMT initial responder and 80.0% (4/5) SOC initial responder could maintain remission.5 FMT initial responder recipients and5SOC initial responder completed 24-months follow up and mainly could maintain remission [FMT vs SOC580% (4/5) vs 60% (3/5)].No adverse events occurred post FMT during long-term follow-up. At Phylum level, *Bacteroidetes, Firmicutes* and *Proteobacteria* were the dominant bacteria of gut microbiota in active UC patients. Compared with donor, the relative abundance of *Bacteroidetes* decreased and *Proteobacteria* increased significantly in active UC patients, *Firmicutes* showed no significant changes. Single fresh FMT could effectively reconstruct the composition of gut microbiota in active UC and maintain stability level with increased *Bacteroidetes* and decreased *Proteobacteria* abundance. FMT significantly reduced the relative abundance of *Escherichia* and increased the relative abundance of *Prevotella* at genera level. Pyruvate metabolism, glyoxylate and dicarboxylate metabolism, pantothenate and CoA biosynthesis showed significantly differences.

**Conclusions:** Single fresh FMT is an effective and safe strategy to induce long-term remission in patients with active UC and could be expected to be an alternative induction therapy for recurrent UC, even primary UC.

**What does this paper add to the literature?:** FMT is an effective and safe therapy for UC. However, long-term efficacy and safety of a single FMT was very limited. The present study found that a single fresh FMT could induce long-term remission in UC with no drugs need and could be expected to be an alternative induction therapy for recurrent UC, even primary UC

## 1. Introduction

Ulcerative colitis (UC), a major subtype of inflammatory bowel disease (IBD), is characterized by chronic recurrent colorectal mucosal inflammation. The exact etiology and pathogenesis of UC remains to be elucidated. Genetic predisposition, deregulation of immunological responses and intestinal barrier dysfunction, dysbiosis of gut microbiota and external environmental triggers had been identified as the pathogenic key factors for UC.IBD has become a global health burden with increasing incidence and prevalence. Globally the incidence and prevalence of UC vary in different regions, which are highest in westernized countries. With the western diet and lifestyle, as well as wide use of antibacterial drugs, social pressure, the incidence of UC is also increasingly prevalent in newly industrialized countries in Africa, South America and Asia including China ^[1-3]^.

The therapeutic goals for IBD are to achieve long-term induction and maintenance of remission and mucosal healing, reduce complications and improve patients’ quality of life. Post-hoc analysis of clinical trials supported the switch of the target from clinical remission to endoscopic healing. Deep remission has been empirically defined as clinical and endoscopic remission ^[4-6]^.Currently, aminosalicylicacid, corticosteroids, immunosuppressants and biological agents are the main drugs to treat UC. Patients with severe disease or serious complications may even require surgical treatment including surgery and endoscopic surgery. However,5-aminosalicylic acids are just suitable for mild to moderate UC and annual relapse rates of up to 25 −40% have been reported despite use of optimal doses of5-ASAs.Corticosteroids play a key role in induction of remission in active UC, but not suitable for maintenance therapy due to experience adverse effects for long-term use and a substantial proportion of steroid-dependent or steroid-resistance patients. Long-term use of immunosuppressants such as thiopurines can cause serious adverse events. For patients with IBD who have failed traditional therapies, Biologics such as infliximab and adalimumab had brought a revolution. However, long-term potential serious adverse events and high costs still limit their clinical application ^[7-9]^.Therefore, new therapeutic strategies need to be constantly explored.

Fecal microbiota transplantation (FMT) refers to transfer gut microbiota from healthy volunteers into patients with gut microbiota dysbiosis related diseases, in order to remodel homeostasis of gut microbiota, achieving the purpose of treatment and even prevention of disease^[10]^.FMT was first recorded in ancient Chinese physician Ge Hong’s “Handbook of Prescriptions for Emergencies” (Eastern Jin Dynasty, AD 317-420). “Drink a liter of healthy fecal juice and you live” was recorded in this famous traditional Chinese medicine works and this method was mainly used to treat patients with food poisoning or severe diarrhea^[11]^. Another famous traditional Chinese works titled “Compendium of Materia Medica” (written by Li Shizhen in the Ming Dynasty) also recorded FMT method. From the perspective of modern medicine, FMT was first reported for the treatment of refractory and recurrent Clostridium infection (RCDI) in 195 8^[12]^. Now FMT is a robust method of manipulation of gut microbiota and act as an extremely effective for RCDI with the efficacy was better than vancomycin via randomized controlled trials (RCTs), FMT has been enrolled clinical treatment guidelines for RCDI therapy in United States and European countries ^[13-14]^.

The first case of UC treated with FMT was reported in 1989^[15]^.Now, due to the efficacy and safety of FMT for RCDI, lots of case reports, case series, andsome RCTs had been reported to investigate the efficacy and safety of FMT for IBD. Our previous research found that FMT is an effective and safe therapy for both paediatric and adult IBD, which might be a potential rescue therapy and even an initial standardized therapy for IBD ^[10]^.Four RCTs have proven the significantly efficacy for inducing remission in active UC ^[16-19]^.UC perhaps represent one of the most robust potential indications for FMT after RCDI ^[20]^.However, there are still no standardized guidelines for treating UC with FMT. The indications for FMT, the route of transplantation, the frequency and dose of FMT, donor selection, and donor stool processing remain controversial. More importantly, data on the long-term efficacy and safety of FMT was very limited. The purpose of the present was to provide the long-term efficacy and safety of single fresh FMT for recurrent active UC. The primary outcome was steroid-free remission of UC, defined as a total Mayo score of 2 with an endoscopic Mayo score of 1or less at week 8. Secondary clinical outcomes included adverse events, quality of life scores and characteristics of gut microbiota before and after FMT treatment. Follow up clinical data were collected for 12 to 24 months.

## 2. Materials and methods

### 2.1 Study design

Patients with recurrent active UC hospitalized at the department of gastroenterology and hepatology, the Second Hospital of Anhui Medical University from April 2017 were enrolled in this study. Patients were randomly divided into single FMT group and standard of care (SOC) group. Patients in FMT group were just treated with single fresh FMT. Patients in SOC group with mild to moderate UC were treated with mesalazine, those with severe UC were given corticosteroids-induced remission, mesalazine maintenance treatment. All participants were 18 years of age or older and gave written informed consents. The ethics committee of the Second Hospital of Anhui Medical University approved the protocols. The trial is registered at Chinese Clinical Trial Registry (ChiCTR2000030080).

### 2.2 Inclusion criteria

Diagnosis of patients with recurrent active UC depended on typical clinical symptoms, endoscopic assessment and histological findings. Mayo scoring system was used to evaluate disease extent. Subjects enrolled in this study should be recurrent active UC with a Mayo score of 4 to 12, which should previously receive a stable dose of5-aminosalicylic acid treatment for at least 4 weeks, but without any other therapy including immunosuppressive agents, biologics and surgery. No history of antibiotics, probiotics or prebiotic use for at least the last month. No history of bowel preparation for at least the last month. No history of FMT treatment. Eligible participants were aged 18-75 years regardless of gender. All patients obtained written informed consent.

### 2.3 Exclusion criteria

Patients with UC are pregnant, have a history of abdominal surgery, have been exposed to antibiotics or probiotics in the last 4 weeks were excluded. Evidences of other infection such as *Clostridium difficile*, cytomegalovirus, epstein-barr virus or extra-intestinal infections requiring antibiotics were excluded. Patients with other comorbidities such as heart, lung, and cerebrovascular disease, history of gastrointestinal malignancy, polyps or IBS, abdominal surgery, or inability to undergo endoscopy were also excluded.

### 2.4 Donor selection

Potential healthy stool donors were sought by a strictly screening questionnaire, sequentially medical interview, and examination followed by blood and stool testing to minimize the risks of disease transmission as previously described ^[25]^. No personal or family history of irritable bowel syndrome, chronic constipation, chronic diarrhea, gastrointestinal cancer, polyps, intestinal tuberculosis and any other chronic gastrointestinal diseases. No personal or family history of diabetes, metabolic syndrome, obesity, hypertension, malnutrition, liver/kidney dysfunction and any other autoimmune or allergic disease such as eczema, asthma and so on. No common detectable enteric pathogens by stool microscopy and culture such as *Entamoeba coli, Clostridium difficile, tuberculosis* and so on. No evidence of infectious diseases such as Epstein-Barrvirus, Cytomegalovirus, hepatitis A, B, C, D and E virus, syphilis, and human immunodeficiency virus (HIV) by blood testing. No history of drug abuse and recent gastrointestinal surgery. No history of antibiotics, chemotherapy drugs, and immunosuppressive agents in the last 3 months. All donors should receive written informed consent from themselves or their guardians.

### 2.5 Donor stool processing

Fresh feces were processed on the morning of the day of FMT. Fresh donor feces need to be processed within 1hour after the donor defecates. In summary, 50 g of freshly passed donor feces were dissolved in 250 ml of sterile 0.9% physiological saline for5minutes with a conventional blender, and then filtered through 2.0 mm, 1.0 mm, 0.5 mm, and 0.25 mm stainless steel filters in order. Finally, the filtered liquid was centrifuged (6000 r / min) at 4 ° C for 15 minutes, and the precipitate was re-dissolved in 150 ml of sterile physiological saline for FMT by colonoscopy.

### 2.6 FMT procedure

Patients in FMT group were not allowed to take antibiotics and aminosalicylic acids like sulfasalazine or mesalamine and a light diet was taken 2-3 days before FMT. Participants received the bowel lavage (polyethylene glycol electrolyte dissolved in 2L water) 4-6 hours prior to FMT. Fresh FMT was processed within 4-6 hours after donor feces processed. In detail, a total of 200 mL of donor fecal slurry was delivered into the right and left colon respectively via colonoscopy. Upon completion of the transplantation, all recipients were required to remain in bed and kept at least for 60 minutes without defecation.

### 2.7 Clinical outcomes and follow-up

The primary endpoint was clinical and mucosal remission at week 8. Clinical remission was defined as Mayo score ≤2with each sub-score≤1, and mucosal remission was defined as Mayo endoscopy sub-score≤1compared with baseline. Clinical response was defined as a decrease in Mayo score ≥30% and ≥3 points when compared with baseline at week 8. Sub-item score of bloody purulent stool decreased ≥ 1score or score 0 or 1was defined as clinical improvement. Patients who achieved clinical response were also enrolled in the clinical response analysis.

The second endpoint was the maintenance of clinical and mucosal remission, and possible adverse events during the long term follow up (12 to 24 months).Patients were followed up at week-2, 4, 8, 12, and 24 and at month-12 to 24 after treatment. Colonoscopy findings and Mayo scores were calculated at week-0 (baseline) and week-4, 8, 24 and month-12after treatment.

Clinical relapse was defined as exacerbation of diarrhea and purulent bloody stool that require drug, including initiation or replacement of drug to induce remission. All patients were followed up by telephone or outpatient service.

### 2.8 Assessment and analysis of gut microbiome

Fresh fecal samples of the donors and pre-FMT and post-FMT treatment to patients were collected using a sterile collection spoon and stored in 3 ml of preservation solution at -°Cfor analysis. Gut microbiota were assessed by 16S ribosomal RNA sequencing. The V4 hyper variable region of the 16s rRNA gene was amplified using the Illumina MiSeq platform high-throughput sequencing and raw sequencing data processed into operational taxonomic units at 97% similarity in stool samples from individual donor and FMT recipients pre and post FMT treatment.

#### 2.8.1 Statistical analyses and visualization

Estimates of alpha diversity were based on an evenly rarefied OUT abundance matrix and included observed richness Observe Species, Shannon, Simpson, ACE, Chao1, using the R packages vegan. And the significance difference of measured alpha-diversity metrics across samples was tested using a nonparametric Kruskal-Wallis rank sum test and Benjamini-Hochbery corrections. Beta diversity of the samples was measured using Bray-Curis distance based on an evenly rarefied OTU abundance table. The β-diversity can estimate the difference in community structure between samples. Statistical differences of measured β-diversity metrics across groups were determined using PERMANOVA with 999 permutations, using adonis in R package vegan. And shared OTU were calculated and visualizing using the R packages Venn diagram. The taxa abundance was measured and plotting using the ggplot2. The LEfSe analysis was performed to identify taxa with differentiating abundance in the different group. LEfSe was an algorithm for high-dimensional biomarker discovery and explanation that identifies genomic features characterizing the differences between two of more biological conditions. Moreover, the indicator analysis based on genera was conducted using R package. Indicator taxa analysis was a way to calculate the probability that any taxon was found in different groups, a taxon with a high indicator value has a high probability of being found within a given treatment and a low probability of being found outside the treatment, and the p-values were corrected with the method of Benjamini-Hochbery using the p.adjust in R. Finally, the results were visualizing using the custom R script based on ggplot2. These analyses were performed using R v3.4.1.

#### 2.8.2 Functional profiling based on bacterial taxonomy

To predict metagenomics function content, the Phylogenetic Investigation of Communities by Reconstruction of Unobserved States (PICRUSt) was used to predict which genes are present using 16s data. The software utilizes a computational approach to predict the functional pathway from the 16s rDNA reads. First, the reads were against a reference collection, Green Genes database, May 2013 version, the closed-reference OTU table was built using qiime. And the resulting OTU table was normalized by normalize_by_copy_number.py. Metagenome predictions were conducted using predict metagenomes.py. And the statistical difference analysis was determined using ANOVA. The results were visualized using custom R script based on ggplot2.

### 2.9 Statistical analysis

An intention-to-treat analysis was done. Baseline demographic, medication and diseases parameters including disease duration, severity and extent are presented using means (SDs) or frequencies (percentages).The categorical variables between groups were compared by Chi-square test. The clinical response in both groups was compared by using Student’s t test. SPSS Statistics v17.0 was used for statistical analysis. Statistical significance was defined as P<0.05.

## 3. Results

### 3.1 Clinical features

26 eligible patients which were diagnosed as recurrent active UC were enrolled in this study, 6 patients were excluded (4 not meeting inclusion criteria, 2 declined to participate). 20 eligible patients a total mayo score of 4 to 12 were enrolled and completed the study. Patients were randomized randomly assigned to single FMT group (n=10) and standard of care group (n=10). FMT group achieved only single fresh FMT via colonoscopy. SOC group achieved mesalazine, achieved corticosteroids if no response to mesalazine. There were no statistically significant differences in baseline demographic and clinical characteristics between the two groups (as shown in table 1). All the enrolled patients had a history of5-ASA treatment, except one patient with5-ASA allergy.

**Table 1.** The Baseline characteristics of study population

| Parameters | FMT group | Control group | *P* value |
| --- | --- | --- | --- |
| Total number | 10 | 10 | - |
| Age(yr), M±SD (range) | 51.5±12.7 (32-70) | 44.6±14.9(22-75) | 0.280 |
| Gender, Female/Male | 8/2 | 8/2 | 1.000 |
| Disease duration (yr) | 5.9±7.3 (0.5-25) | 6.9±9.3(0.3-30) | 0.777 |
| Disease severity |  |  | 0.895 |
| Severe % (n) | 50 (5/10) | 40(4/10) | - |
| Moderate % (n) | 40 (4/10) | 50(5/10) | - |
| Mild % (n) | 10 (1/10) | 10(1/10) | - |
| Disease extent |  |  | 1.000 |
| Pancolitis % (n) | 50 (5/10) | 50 (5/10) | - |
| Left-sided colitis % (n) | 50 (5/10) | 50 (5/10) | - |
| Concomitant medications |  |  | - |
| Mesalazine | 90 (9/10) | 100 (10/10) | - |
| Allergy to 5-ASA | 10(1/10) | - | - |
| Total Mayo score | 9.5±2.5 | 8.6±2.9 | 0.462 |
| Endoscopic Mayo score | 2.4±0.7 | 1.9±0.7 | 0.137 |

### 3.2 Primary outcome

In FMT group, all patients (n=10)received FMT therapy,90% (9/10) UC patients achieved clinical symptom improvement within 2 weeks after FMT. Clinical symptoms including purulent bloody stool, defecation frequency, abdominal pain and abdominal discomfort were significantly improved. Compared with baseline, purulent bloody stool and defecation frequency were significant decreased. Mean abdominal pain score was significantly decreased from baseline value 4.5 ±2.2 to 0.9±1.6, and mean diarrheal frequency was significantly decreased from baseline value 8.8±3.8 to 2.5 ±2.7 two weeks after FMT (as shown in Figure1a and Figure 1b). According to Mayo score system, 90% FMT recipients achieved primary endpoint at week-8. Compared with base line, FMT significantly induced clinical remission (*P*=0.000).

**Figure 1.**
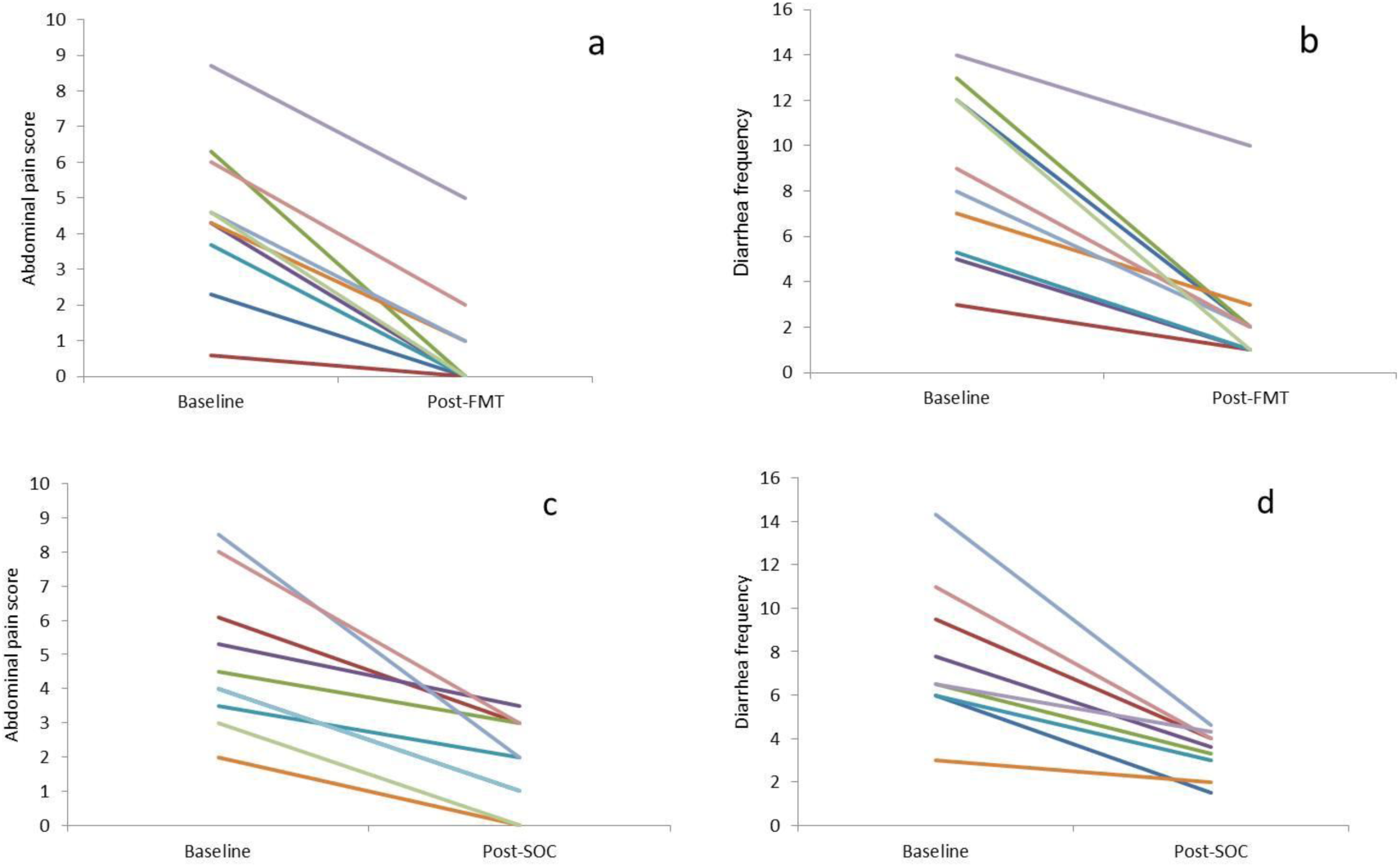
Clinical response to single FMT (a, b) and stander of care (c, d) after two weeks of treatment. **a:** Abdominal pain scores of patients with active UC at baseline(4.5 ±2.2) and 2 weeks after single FMT(0.9±1.6) (n=10). **b:** Diarrheal frequency of patients with active UC at base line (8.8±3.8) and 2 weeks after single FMT (2.5 ±2.7), **c:** Abdominal pain scores of patients with active UC at baseline (4.9±2.1) and 2 weeks after SOC therapy (1.8±1.3) (n=10), **d:** Diarrheal frequency of patients with active UC at baseline (7.8±3.1) and 2 weeks after SOC therapy (3.3±1.0).FMT5 Fecal microbiota transplantation, SOC=stander of care.

In SOC group (n=10),50% (5 /10) UC patients achieved clinical symptom improvement within 2 weeks. Mean abdominal pain score was significantly decreased from baseline value 4.9±2.1to 1.8±1.3, and mean diarrheal frequency was significantly decreased from baseline value 7.8±3.1to 3.3±1.0 two weeks after SOC therapy (as shown in figure1c and figure 1d).50% (5 /10) patients in SOC group achieved primary endpoint at week-8. Compared with SOC group, FMT could achieve clinical remission more quickly and effectively at week-8 (*P*=0.019, as shown in figure 2a and figure 2b).

**Figure 2.**
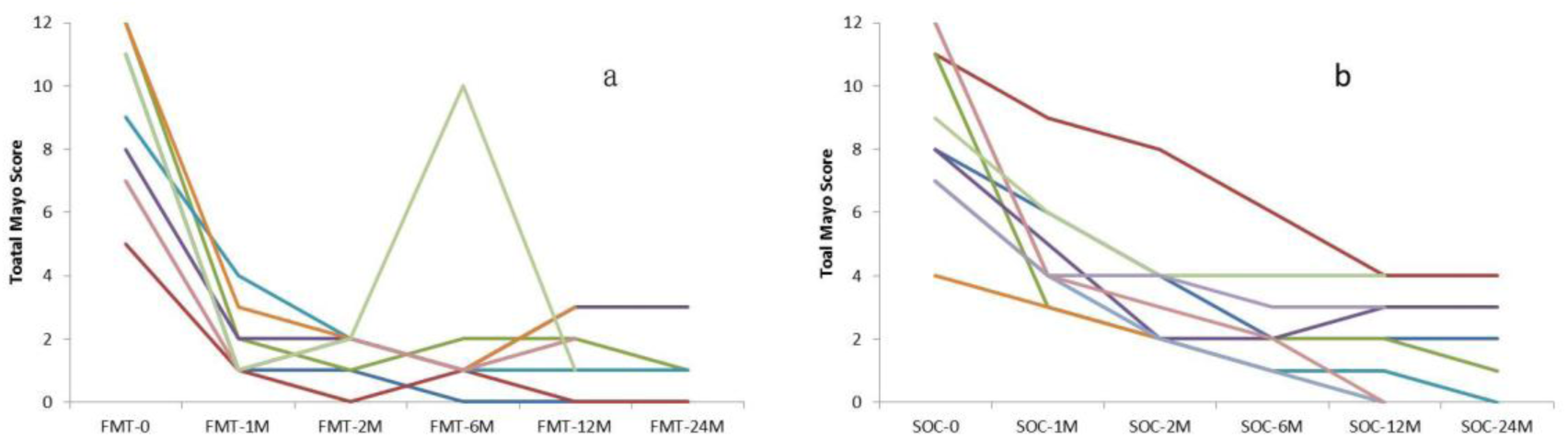
Mayo scores of long-term follow up of single FMT (a) and stander of care (b) to active UC. Assessed at week 8, FMT recipients Mayo scores significantly lower than that of SOC recipients (*P*=0.019). Reassessed at 12-month, there were no significant difference between two groups (*P*=0.691). FMT5 Fecal microbiota transplantation, SOC=stander of care.

### 3.3 Clinical outcomes of long-term follow-up

In FMT group, one patient who initially responded to FMT maintained remission for 6 months and relapsed. The patient received the same donor FMT via colonoscopy again. Unfortunately, he did not response to the second FMT treatment and transferred to corticosteroids induction plus mesalazine maintenance therapy. This patient maintained clinical and mucosal remission during the following long-term follow up. The rest 8 initial FMT respondents were reassessed at month-12 after FMT treatment, 75 % (6/8) maintained clinical and mucosal remission with no drug required and no adverse events.5patients who initially responded to FMT completed 24 months follow-up, 4 patients maintained clinical and mucosal remission with no drug required and no adverse events, 1patients relapsed with mayo score 3 and mesalazine was chosen as rescue therapy. Figure 3 was the colonoscopy performance pre and post FMT of chronic severe recurrent active UC with Mayo score 12 at baseline. She was 36 years old and allergic to mesalazine. She maintained clinical and mucosal remission for 24 months after single fresh FMT.

**Figure 3.**
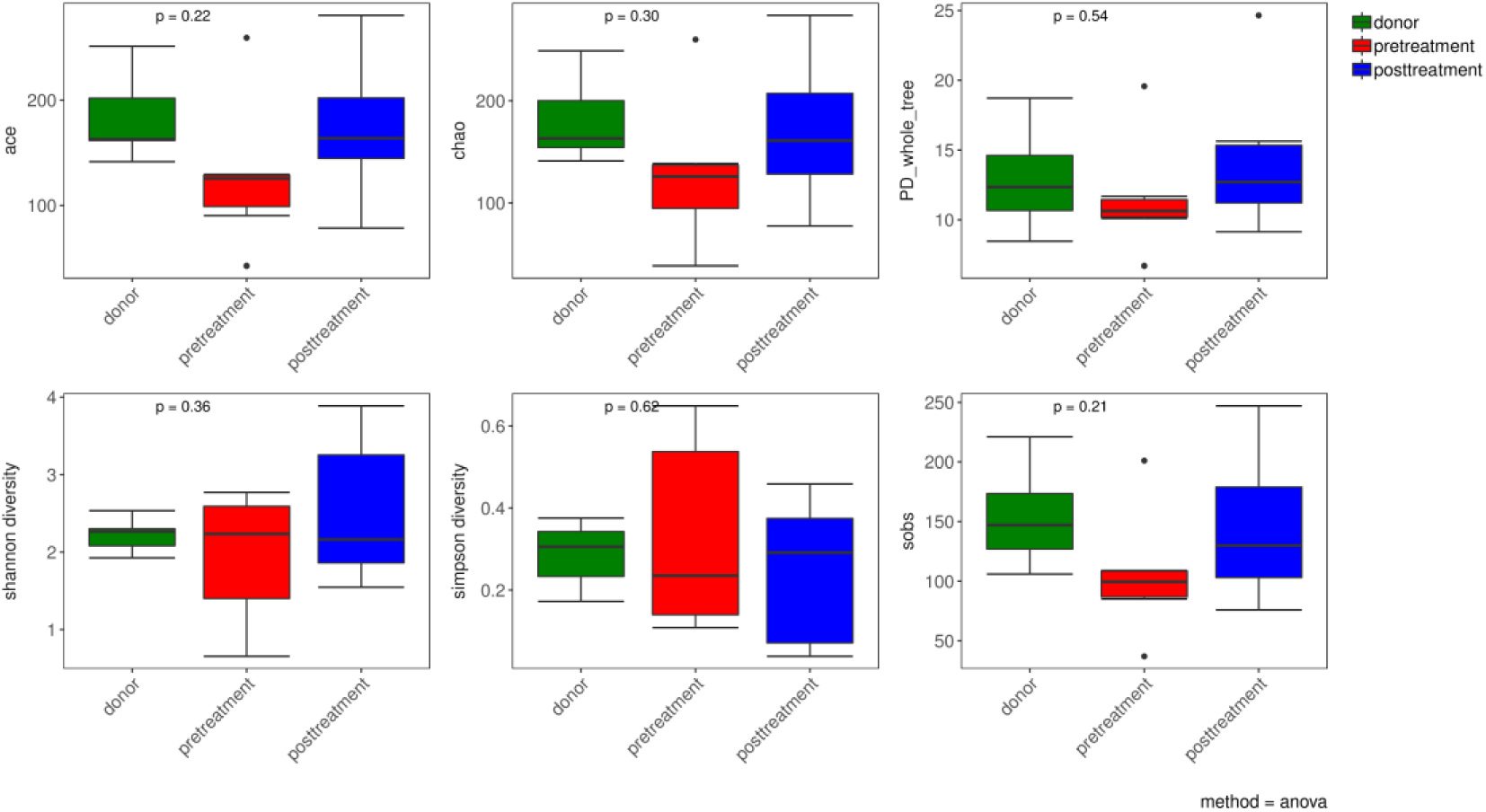
Alpha diversity index box chart. The abscissa represents sample grouping and the ordinate was alpha index, healthy donors, active UC patients (marked as pretreatment) and post FMT treatment (marked as post-treatment) showed no significant difference (*p*> 0.05).

In SOC group,5initial SOC respondents were reassessed at 12-month after therapy, 80% (4/5) patients maintained clinical and mucosal remission.5 SOC therapy patients completed 24-months follow up, 3 patients maintained clinical and mucosal remission,2 patients relapsed with mayo score 3 and 4 respectively. There was no significant difference reassessed at 12-month between FMT and SOC group (P=0.691, as shown in figure 2).

### 3.4 Safety of FMT

Adverse events were recorded during FMT and long-term follow-up (12-24 months). All patients could tolerate FMT treatment, and no serious adverse events occurred during FMT treatment. Some patients developed mild abdominal pain, bloating, nausea, but all resolved spontaneously within 24 hours after FMT treatment. One patient developed diarrhea after treatment and relieved within 24 hours without any medical intervention. A58-year-old female UC patient in FMT group achieved clinical remission and discharged1week after FMT. However, she suffered from aggravated purulent bloody stool, defecation frequency abdominal pain and fever two weeks later after FMT due to epstein-barr virus infection. After antiviral therapy for5days, the patient achieved clinical remission again and was able to maintain remission during the 2 years of follow-up. However, no evidence of acute epstein-barr virus infection was detected in this patient before FMT or in the donor of this patient.

Long term safety: Although many studies have reported adverse reactions such as fever, abdominal pain, bloating, diarrhea after FMT, but most are self-limiting. And no side reactions were observed and no infection of certain pathogens according the 12 to 24 months long-term following up. No patients suffered from other chronic diseases such as immune system diseases, non-alcoholic fatty liver disease. All patients had good tolerance to FMT treatment.

### 3.5 Assessment and Analysis of gut microbiome

According to the rarefaction curves plateau with the current sequencing indicating that most of the diversity has already been captured in all samples. Alpha diversity index calculation by abundance index (Chao1and ACE), diversity index (shannon and simpson). The alpha diversity index of the fecal microbiota in active UC patients, healthy donors and post FMT treatment showed no significant difference (As shown in Figure 3, *p*> 0.05).To measure the level of similarity between gut microbial communities, Analysis of similarities (ANOSIM) was performed. The data revealed an apparent separation in the structure of the gut microbiota in each group (As shown in Figure 4, *p*=0.011).

**Figure 4.**
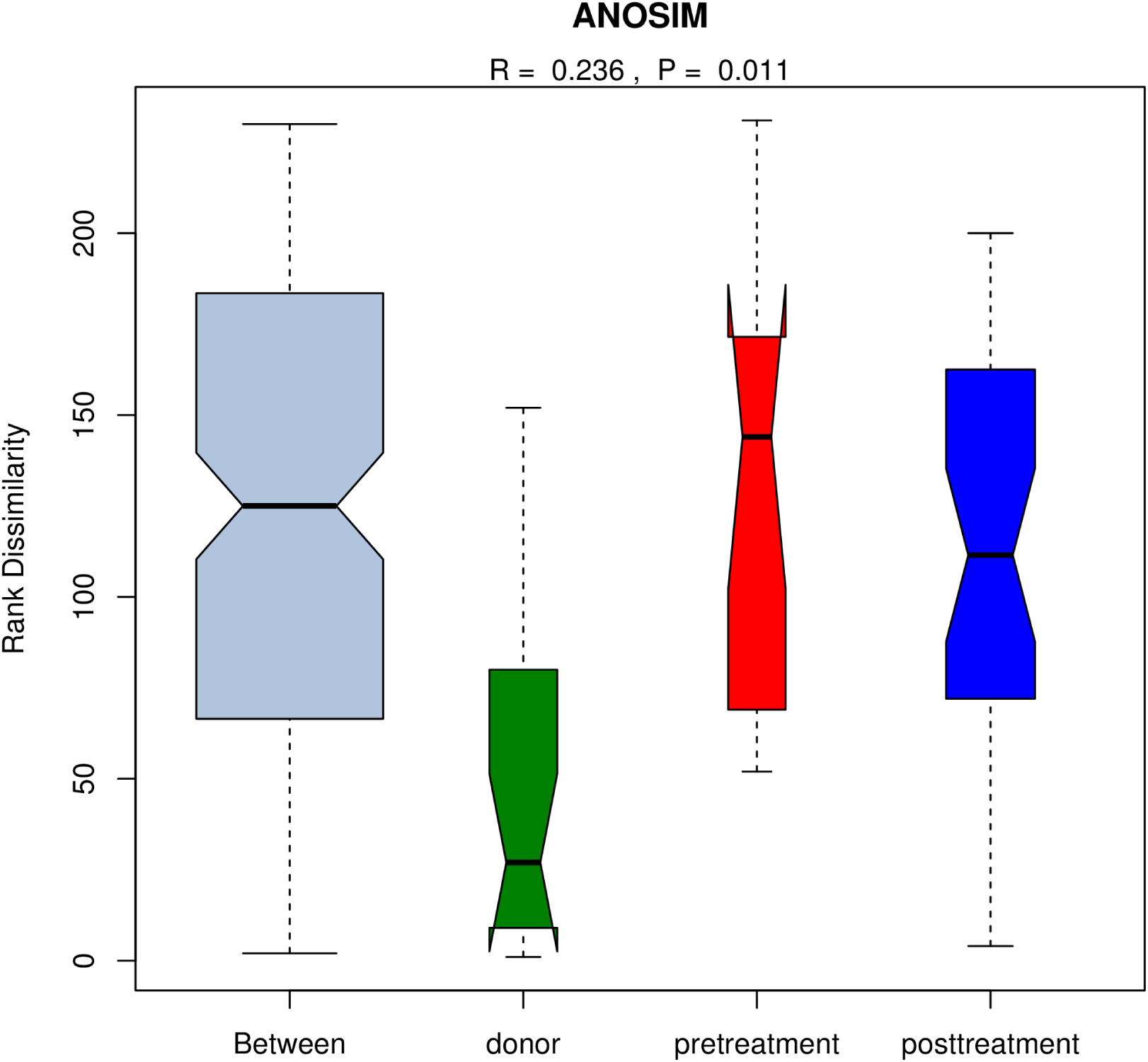
Analysis of similarities (ANOSIM)

PCA and principal coordinate analysis (PCoA) was used to indicate the similarity of microbiota composition among samples. PCoA analyses revealed that the gut microbiota in UC patients significantly deviated from the healthy donors. Treatment of FMT could improve the distance markedly and clustered tightly together, showed a trend similar to their related donors, but not return to the level of e healthy donors (as shown in Figure5). The system clustering tree also indicated a significant difference existed in between UC patients and healthy donors.

Subsequently, linear discriminant analysis effect size (LEfSe) was used to identify differential microorganism communities between groups. The taxonomic profiles showed that the phylum *Bacteroidetes, Firmicutes* and *Proteobacteria* were dominant bacteria of fecal microbiotain healthy donors and active UC patients. The relative abundance of *Bacteroidetes* was significantly decreased and *Proteobacteria* was significantly increased in active UC patients. *Firmicutes* showed no significant changes among healthy donors and active UC patients. Compared with healthy donors, patients with active UC showed increased ratio of *Firmicutes* and *Bacteroidetes*. Single fresh FMT could significantly reconstruct the dysbiosis of gut microbiota and maintain stability level with increased portion of *Bacteroidetes* and decreased portion *of Proteobacteria*.

At the genera level, some specific bacterial biomarkers were identified. The relative abundance of *Escherichia* was significantly increased in active UC patients, which was significantly decreased after FMT. High abundance of *Prevotella* was found in donors gut. FMT-treated patients who achieved remission also tended to have higher abundance of *Prevotella* (as shown in Figure 6 and Figure 7).

**Figure 5.**
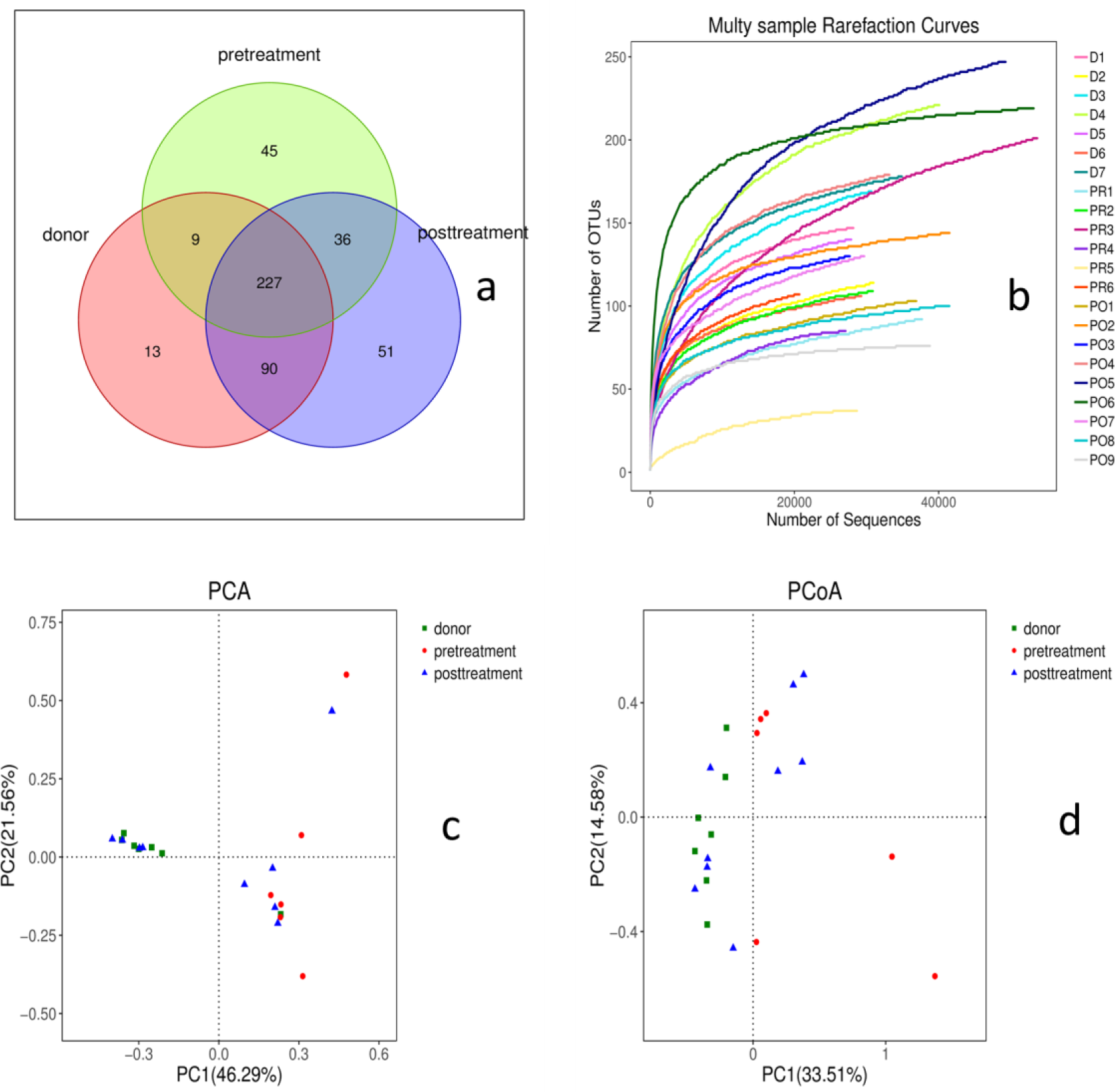
Beta diversity index box chart. Principal coordinate analysis (PCoA) of gut microbiota among healthy donors, active UC and post FMT treatment, the distance between the samples represents the similarity of gut microbiota composition, a closer distance indicates higher similarity. Abbreviations: Patient5 active UC patients, FMT5 post-FMT treatment, Donor5healthy donors

**Figure 6.**
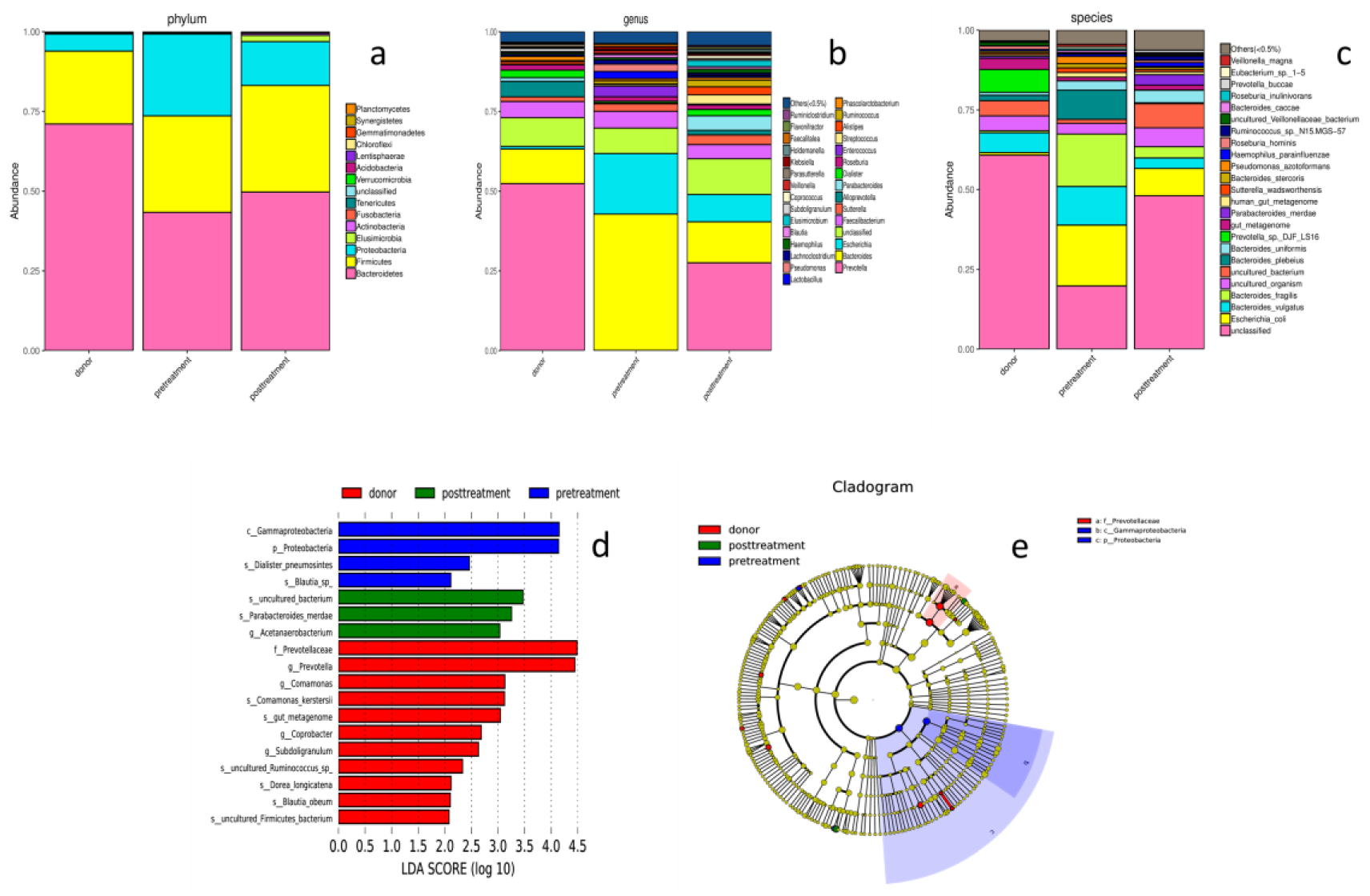
Histogram of taxonomic profiles of gut microbiota among healthy donors, active UC (marked as pretreatment) and post FMT treatment (marked as post-treatment) at the phylum (a), genera (b)and species (c) level. LDA score (d) and Cladogram (e). *Prevotella* was dominant genera of gut microbiotain healthy donors, relative high abundance of *Prevotella* increased after FMT treatment in UC patients.

**Figure 7.**
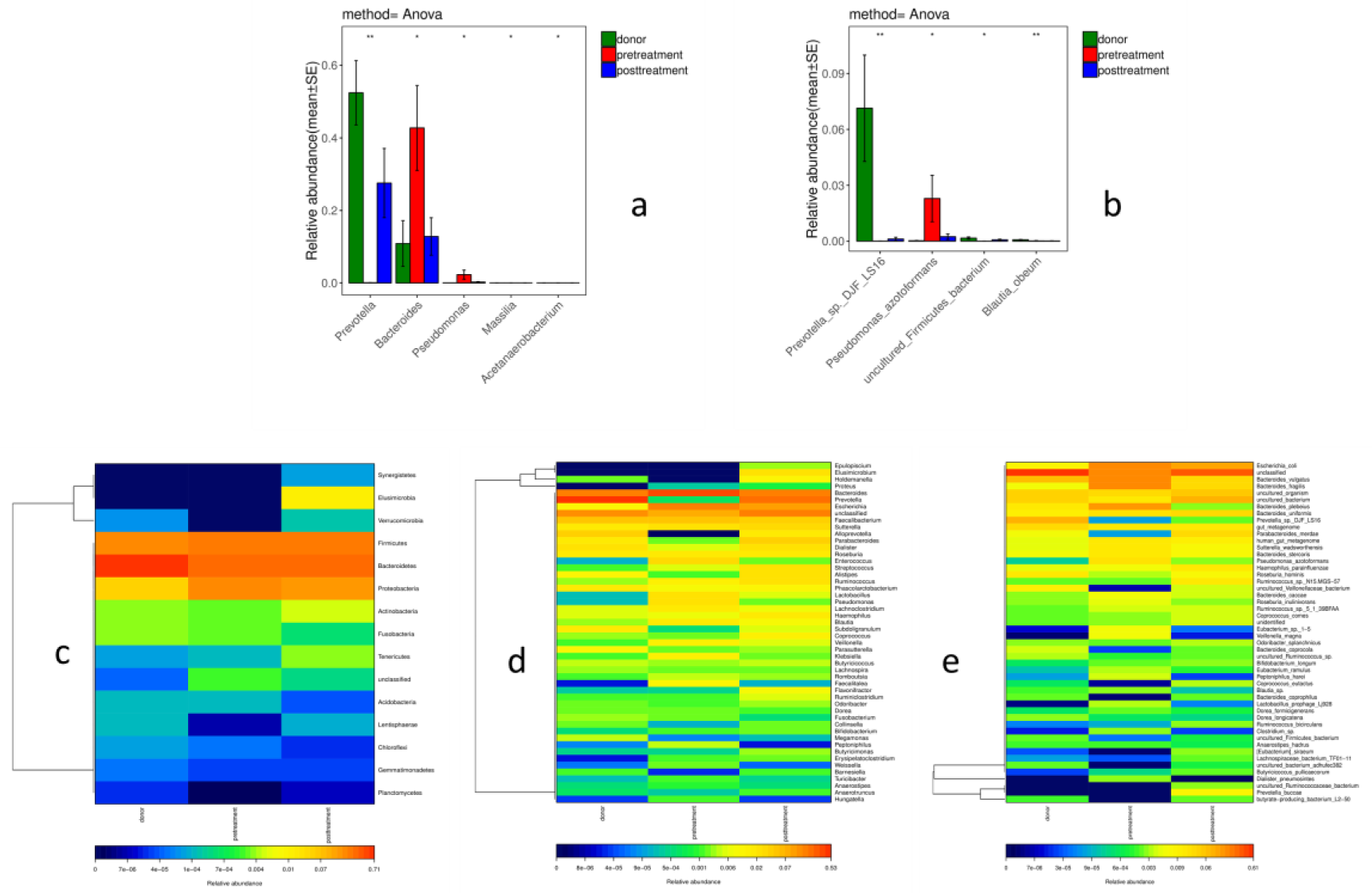
Heat map-raim bow of taxonomic profiles of gut microbiota among healthy donors, active UC (marked as pretreatment) and post FMT treatment (marked as post-treatment) at the phylum (c), genera (d) and species (e) level. The relative high abundance of bacteria at genera (a) and species (b) level

PICRUSt tool was used to predict the functional profiles of gut microbiota with the predicted metagenome, Kyoto Encyclopedia of Genes and Genomes (KEGG) pathway functions were categorized using the PICRUSt. The relative abundances of pyruvate metabolism, sulfur metabolism, pantothenate and CoA biosynthesis, glyoxylate and dicarboxylate metabolism, synthesis and degradation of ketone bodies and other transporters were significantly different (as shown in figure 8).

**Figure 8.**
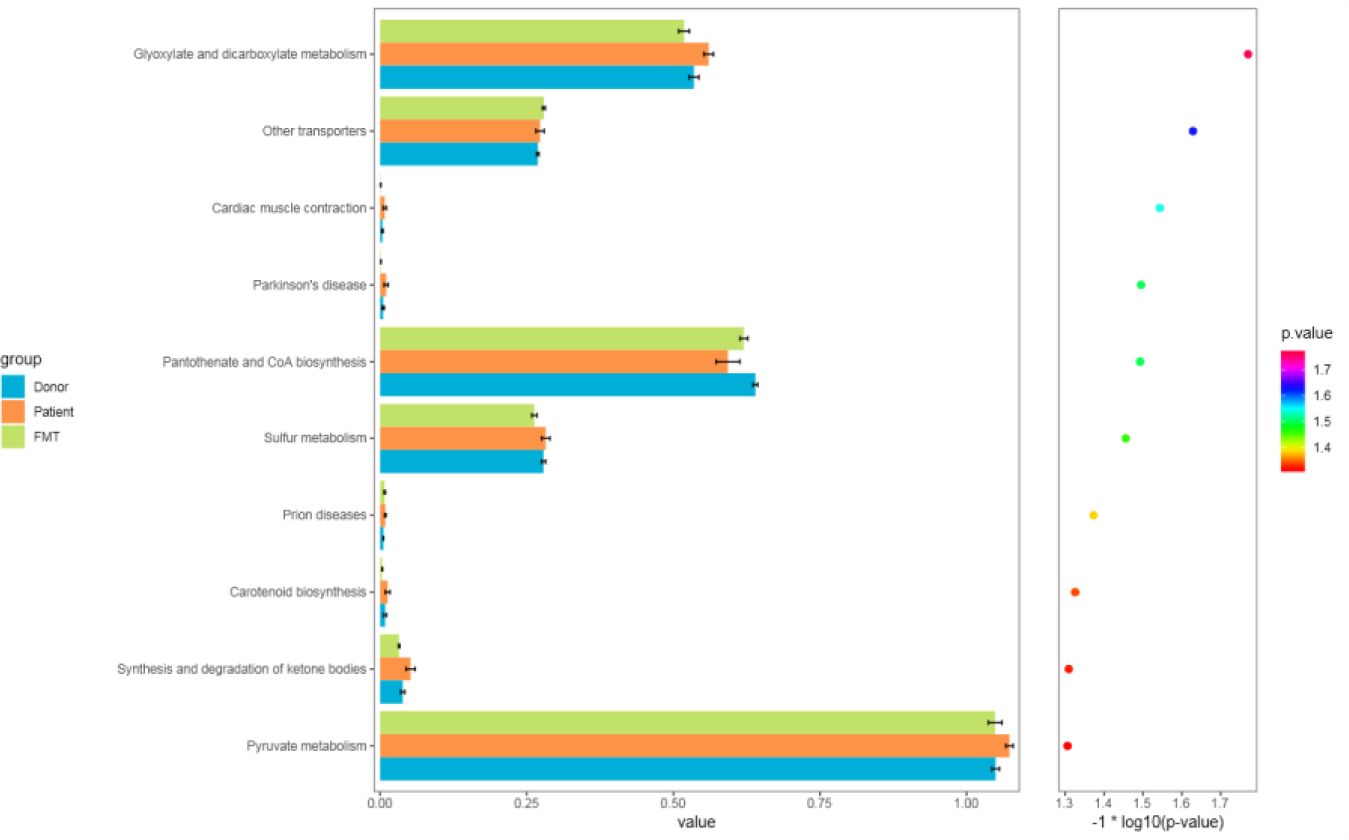
PICRUSt predicted the distribution of KEGG secondary metabolic pathways.

## 4. Discussion

FMT is a robust method to increase the diversity of recipient gut microbiota. Engraftment of donor microbiota resulted in a long-lasting response in patients with RCDI ^[21]^.Growing evidences confirmed that gut microbiota of patient with UC is characterized by lower diversity and different proportion of certain microorganisms. Manipulation of gut microbiota via FMT is an emerging novel potential promising therapy for IBD ^[22-23]^.However, due to the complex pathogenesis of UC and multiple influencing factors, the optimum intensity and duration of FMT have not been defined so far.

### 4.1 Frequency factor influence the outcomes of FMT

One RCT data showed that multi-donor intensive FMT induced clinical remission and endoscopic improvement in active UC, which was associated with distinct microbial changes that relate to outcome ^[18]^. The other RCT also showed that pooled FMT resulted in a higher likelihood of remission in adult patients with mild to moderate active UC. The diversity of gut microbiota in patients receiving pooled FMT was more abundant than that of a single donor, but it was uncertain to better efficacy or not ^[19]^. Multi-session FMT could induce clinical remission and aid in steroid withdrawal for patients with steroid-dependent UC ^[24]^. For UC patients with remission by multi-session FMT, stable dose of SOC (5 -ASA with/without azathioprine) plus continuous FMT treatment presented a higher tendency to maintain a steroid-free clinical remission and significantly superior to placebo in both endoscopic and histological remission ^[25]^. In view of the above data, multi-session FMT seems to be an effective therapy for maintaining remission in patients with UC. Nevertheless, an open-label pilot study reported that daily oral multi-donor FMT capsules for fifty days as a supplement to SOC, the symptoms and health related life quality of UC only temporarily improved ^[26]^.

In our study, FMT recipient just achieved treatment with single fresh FMT, 90% could achieve clinical symptom improvement within 2 weeks and could achieve primary endpoint at week-8. Compared with SOC group, FMT could achieve clinical remission more quickly and effectively at week-8, particularly for moderately to severely active UC, which is consistent with the previously reported meta-analysis ^[10]^. These may be correlated with different degrees of dysbiosis of gut microbiota in UC, and it is expected that the inflammatory areas are more serious than the non-inflammatory areas ^[27-28]^.Of course, the possible effects of other influencing factors such as donor, gut microbiota characteristics of donors and UC patients on the efficacy of FMT were also discussed later.

### 4.2 Donor factors influence the outcomes of FMT

To date, there have been no randomized studies comparing differences in efficacy, related or unrelated donors should both be considered acceptable, there were clear advantages to using FMT from a stool bank, from a healthy unrelated, in particular with regards to monitoring and traceability ^[29]^. Based on previously reported data including clinical guidelines, the donors most aged more than 18 years. According to traditional Chinese medicine theory, young healthy volunteers and even children were more suitable as donors. One RCT study reported a higher treatment success with one particular donor compared with the other donors ^[16]^.Donor selection may play much more important role in the therapy of ulcerative colitis than that in the scenario for CDI ^[30]^. In the present study, according to the donor selection criteria, 10 eligible donors (7 female, 3 male) were selected. Age ranged from5to 20 years old, mean age was 11.3±4.4 years old. 90% donors were the first-degree relatives of the patient with UC. In this study, UC patients were able to obtain significant clinical remission from a single fresh FMT, which may be correlated to the selection of young donors. This may be an interesting topic for further study, especially in the RCT.

### 4.3 Role of reconstruction of gut microbiota via FMT

Through assessment and analysis of gut microbiome in UC and healthy donor, we found that the diversity and richness of gut microbiota of active UC patients was significantly different from that of healthy donors. The diversity was reduced and the relative abundance of *Bacteroides* was decreased, *Proteobacteria* was increased significantly, but *Firmicutes* showed no significant change. Dysbiosis of gut microbiota in active UC patient may be reconstructed by FMT, which was similar to that of the donor. At genus level, the relative abundance of *Escherichia* was decreased and the level of *Prevotella* was markedly increased after FMT. Phyla *Bacteroidetes* are mainly composed of *Bacteroides* and *Prevotella* genera, which are thought to share a common ancestor. *Bacteroides* is the predominant bacteriain human gut with western diet style characterized by high protein and animal fats, while in non-westernized populations consuming plant-rich diet, *Prevotella* dominates in gut microbiata ^[31]^. *Prevotella* is a large genus with high species diversity and high genetic diversity among strains, which makes it difficult to predict their function with obvious individual difference^[32]^.The role of *Prevotella* in the pathogenesis of UC is still controversial. One single species isolate *Prevotellacopr i*CB7 has been used for different studies and can be beneficial or detrimental, depending on the context ^[32]^. Intriguing, some papers suggested that *Prevotella* might be regarded as a beneficial bacterium, but not a pathobiont. For example, the enrichment of *Prevotella copri* was observed in healthy individuals taking fiber-rich diet that normally exhibit anti-inflammatory activities. The most consistent finding is that *Escherichia*, specifically *Escherichia coli. E. coli* was increased in IBD. However, due to technical limitations, the vast majority of microbiota studies in IBD do not analyze the microbiota at the strain level. So it is unknown whether the increase or decrease of certain microbiota is due to pathogenic strains or “protective” strains ^[33]^. An individual’s response to FMT may predominantly depend on the capability of the donor’s microbiota to engraft and reverse the microbial community dysbiosis associated with a specific disease phenotype; these factors need to be investigated in future studies ^[34]^.

### 4.4 long-term efficacy and safety of single FMT in UC patients

Borody et al ^[35]^ reported 6 patients with UC received FMT enema for five consecutive days, all patients obtain complete reversal of symptoms and maintain remission for 1-13 years with no drugs required. Ding ^[36]^ reported that through follow-up of 1-5 years, step-up FMT was safe and effective for patients with moderate to severe UC and improved the quality of life. But this step-up FMT strategy means repeated FMT, step1was single FMT, step2 was multiple FMT (at least 2 times), then one or more FMT in combination with corticosteroids or cyclosporine if no response to step 1or 2. In our study, FMT recipient just achieved treatment with single fresh FMT. During the long-term follow-up, most of initial FMT respondents were able to maintain mucosal remission with no drug required and no obvious adverse events. We speculate that various factors could be attributed to the positive outcomes achieved in our study. Firstly, fresh fecal slurry contains more beneficial microorganisms and metabolites. The fresh transplant strategy within 6 hours reduced the loss of beneficial microorganisms. Furthermore, the whole colorectal instillation of fresh fecal slurry used a colonoscopic route ensured good volumes of donor’s fecal slurry. All FMT recipients were required to remain in bed and kept at least for 60 minutes without defecation, which ensured a good retention time. The role of bowel preparation in published literature is controversial. Some views hold that an adequate pre-FMT bowel preparation may help in clearing the pro-inflammatory bacteria and colonization of donor microbiota in recipient gut ^[37]^.However, there are also views that bowel preparation with polyethylene glycol may itself cause changes in microbiota, thus produce results not attributable to FMT ^[38]^.Of course, this is a small sample study and perhaps the effective ratio may change or even decrease with the increase of sample size. Of cause, large sample RCT studies are needed.

## 5. Summary

Single fresh FMT is an effective and safe strategy to induce long-term remission in patients with active UC. Single fresh FMT could effectively reconstruct the composition of gut microbiota in active UC. FMT is expected to be an alternative induction therapy for recurrent UC, even primary UC. Although this was a small sample study, it still could provide reference for treating active UC, especially recurrent UC due to the long-term follow-up.

## Data Availability

All data referred to in the manuscript are availabile

## References

1. Ng SC, Shi HY, Hamidi N, Underwood FE, Tang W, Benchimol EI, Panaccione R, Ghosh S, Wu JCY, Chan FKL, Sung JJY, Kaplan GG. Worldwide incidence and prevalence of inflammatory bowel disease in the 21st century: a systematic review of population-based studies. Lancet. 2018 Dec 23; 390(10114):2769–2778.

2. Molodecky NA, Soon IS, Rabi DM, Ghali WA, Ferris M, Chernoff G, Benchimol EI, Panaccione R, Ghosh S, Barkema HW, Kaplan GG. Increasing incidence and prevalence of the inflammatory bowel diseases with time, based on systematic review. Gastroenterology (2012) 142(1):46-54.e42

3. da Silva BC, Lyra AC, Rocha R, Santana GO. Epidemiology, demographic characteristics and prognostic predictors of ulcerative colitis World J. Gastroenterol., 20 (2014): 9458–9467.

4. Nishida A, Inoue R, Inatomi O, Bamba S, Naito Y, Andoh A. Gut microbiota in the pathogenesis of inflammatory bowel disease. Clin J Gastroenterol. 2018;11(1):1–10

5. ZuoT, NgSC. The gut microbiota in the pathogenesis and therapeutics of inflammatory bowel disease Front Microbiol, 2018,25;9:2247.

6. Cohen LJ, Cho JH, Gevers D, Chu H. Genetic factors and the intestinal microbiome guide development of microbe-based therapies for inflammatory bowel diseases. Gastroenterology. 2019 Jun;156(8):2174–2189.

7. Lemaitre M, Kirchgesner J, Rudnichi A, Carrat F, Zureik M, Carbonnel F, Dray-Spira R. Association between use of thiopurines or tumor necrosis factor antagonists alone or in combination and risk of lymphoma in patients with inflammatory bowel disease JAMA. 2017 Nov 7;318(17):1679–1686.

8. Bemelman WA1; S-ECCO collaborators. Evolving role of IBD surgery J Crohns Colitis, 2018 Jul 30;12(8):1005–1007.

9. Shen B. Interventional IBD: The Role of Endoscopist in the Multidisciplinary Team Management of IBD. Inflamm Bowel Dis. 2018 Jan 18;24(2):298–309

10. Fang H, Fu L, Wang J. Protocol for Fecal Microbiota Transplantation in Inflammatory Bowel Disease: A Systematic Review and Meta-Analysis. Biomed Res Int. 2018 Sep 13;2018:8941340

11. Zhang F, Luo W, Shi Y, Fan Z, Ji G. Should we standardize the 1,700-year-old fecal microbiota transplantation? Am J Gastroenterol.2012,107(11):1755–1755.

12. Eiseman B, Silen W, Bascom G S, Kauvar A J. Fecal enema as an adjunct in the treatment of pseudomembranous. Surgery. 1958;44(5):854–59.

13. Surawicz C M, Brandt L J, Binion D G, et al. Guidelines for diagnosis, treatment, and prevention of Clostridium difficile infections. Am J Gastroenterol. 2013 Apr;108(4):478–98.

14. Debast S B, Bauer M P, Kuijper E J, et al. European society of clinical microbiology and infectious diseases: update of the treatment guidance document for *Clostridium difficile* infection. Clin Microbiol Infect. 2014 Mar;20 Suppl 2:1–26.

15. Bennet J D, Brinkman M. Treatment of ulcerative colitis by implantation of normal colonic flora. The Lancet, 1989,1 (8630):164.

16. Moayyedi P, Surette MG, Kim PT, et al. Fecal microbiota transplantation induces remission in patients with active ulcerative colitis in a randomized controlled trial. Gastroenterology 2015;149:102–109

17. Rossen NG, Fuentes S, van der Spek MJ, et al. Findings from a randomized controlled trial of fecal transplantation for patients with ulcerative colitis. Gastroenterology 2015;149:110–118.

18. Paramsothy S, Kamm MA, Kaakoush NO, Walsh AJ, van den Bogaerde J, Samuel D, Leong RWL, Connor S, Ng W, Paramsothy R, Xuan W, Lin E, Mitchell HM, Borody TJ. Multidonor intensive faecal microbiota transplantation for active ulcerative colitis: a randomized placebo-controlled trial. Lancet 2017;389:1218–1228.

19. Costello SP, Hughes PA, Waters O, et al. Effect of fecal microbiota transplantation on 8-week remission in patients with ulcerative colitis: a randomized clinical trial. JAMA 2019;321:156–64.

20. Colman RJ, Rubin DT. Fecal microbiota transplantation as therapy for inflammatory bowel disease: a systematic review and meta-analysis. J Crohn Colitis 2014;8:1569e81.

21. Weingarden A, González A, Vázquez-Baeza Y, Weiss S, Humphry G, Berg-Lyons D, Knights D, Unno T, Bobr A, Kang J, Khoruts A, Knight R, Sadowsky MJ. Dynamic changes in short- and long-term bacterial composition following fecal microbiota transplantation for recurrent Clostridium difficile infection Microbiome 2015; 3:10

22. Anderson JL, Edney RJ, Whelan K. Systematic review: faecal microbiota transplantation in the management of inflammatory bowel disease. Aliment Pharmacol Ther, 2012; 36:503e16.

23. Cochrane Database of Systematic Reviews Fecal transplantation for treatment of inflammatory bowel disease Cochrane Systematic Review - Intervention Version published: 13 November 2018.

24. Sood A, Mahajan R, Juyal G, Midha V, Grewal CS, Mehta V, Singh A, Joshi MC, Narang V, Kaur K, Sidhu H. Efficacy of fecal microbiota therapy in steroid dependent ulcerative colitis: a real world intention-to-treat analysis. Intest Res. 2019 Jan;17(1):78–86.

25. Sood A, Mahajan R, Singh A, Midha V, Mehta V, Narang V, Singh T, Singh Pannu A. Role of Fecal Microbiota Transplantation for maintenance of remission in patients with ulcerative colitis:a pilot study. J Crohns Colitis, 2019;13(10):1311–1317.

26. Cold F, Browne PD, Günther S, Halkjaer SI, Petersen AM, Al-Gibouri Z, Hansen LH, Christensen AH. Multidonor FMT capsules improve symptoms and decrease fecal calprotectin in ulcerative colitis patients while treated-an open-label pilot study. Scand J Gastroenterol. 2019 Mar;54(3):289–296.

27. Walker AW, Sanderson JD, Churcher C, et al. High-throughput clone library analysis of the mucosa-associated microbiota reveals dysbiosis and differences between inflamed and non-inflamed regions of the intestine ininflammatory bowel disease. BMC Microbiol, 2011;11:7.

28. Sepehri S, Kotlowski R, Bernstein C N, and Krause D O. Microbialdiversity of inflamed and noninflamed gut biopsy tissues in inflammatory bowel disease. Inflamm. Bowel Dis. 2007,13: 675–683.

29. Mullish BH, Quraishi MN, Segal JP, McCune VL, Baxter M, Marsden GL, Moore DJ, Colville A, Bhala N, Iqbal TH, Settle C, Kontkowski G, Hart AL, Hawkey PM, Goldenberg SD, Williams HRT. The use of fecal microbiota transplant as treatment for recurrent or refractory Clostridium difficile infection and other potential indications: joint British Society of Gastroenterology (BSG) and Healthcare Infection Society (HIS) guidelines. Gut 2018; 67(11):1920–1941

30. Allegretti JR, Mullish BH, Kelly C, Fischer M. The evolution of the use of faecal microbiota transplantation and emerging therapeutic indications. Lancet 2019; 394 (10196):420–431.

31. Wu GD, Chen J, Hoffmann C, Bittinger K, Chen YY, Keilbaugh SA, Bewtra M, Knights D, Walters WA, Knight R, Sinha R, Gilroy E, Gupta K, Baldassano R, Nessel L, Li H, Bushman FD, Lewis JD. Linking long-term dietary patterns with gut microbial enterotypes Science, 2011 Oct 7;334(6052):105–8.

32. Ley RE. Gut microbiota in 2015: Prevotella in the gut: choose carefully. Nat Rev Gastroenterol Hepatol. 2016 Feb;13(2):69–70.

33. Pittayanon R, Lau JT, Leontiadis GI, Tse F, Yuan Y, Surette M, Moayyedi P. Differences in Gut Microbiota in Patients With vs Without Inflammatory Bowel Diseases: A Systematic Review. Gastroenterology. 2019 Dec 5.pii: S0016-5085(19)41893-3.

34. Ng SC, Kamm MA, Yeoh YK, Chan PKS, Zuo T 2, Tang W, Sood A, Andoh A, Ohmiya N, Zhou Y, Ooi CJ, Mahachai V 11, Wu CY, Zhang F, Sugano K, Chan FKL Scientific frontiers in faecal microbiota transplantation: joint document of Asia-Pacific Association of Gastroenterology (APAGE) and Asia-Pacific Society for Digestive Endoscopy (APSDE). Gut. 2020;69 (1):83–91.

35. Borody TJ,Warren EF,Leis S, et al. Treatment of ulcerative colitis using fecal bacterio therapy J Clin Gastroenterol. 2003, 37(1):42–47.

36. Ding X, Li Q, Li P, Zhang T, Cui B, Ji G, Lu X, Zhang F. Long-Term Safety and Efficacy of Fecal Microbiota Transplant in Active Ulcerative Colitis. Drug Saf 2019 Jul; 42(7):869–880.

37. Agrawal M, Aroniadis OC, Brandt LJ, et al. The long-term efficacy andsafety of fecal microbiota transplant for recurrent, severe, and complicatedclostridium difficile infection in 146 elderly individuals J Clin Gastroenterol 2016;50:403–7.

38. Jalanka J, Salonen A, Saloj.rvi J, et al. Effects of bowel cleansing on the intestinal microbiota Gut 2015;64:1562–8.

